# Association between central blood pressure, arterial stiffness and low cognitive scores in South African adults

**DOI:** 10.1101/2022.03.29.22273099

**Authors:** Feziwe Mpondo, Ashleigh Craig, Andrea Kolkenbeck-Ruh, Larske Soepnel, Simone Crouch, Sanushka Naidoo, Shane A. Norris, Justine Davies, Lisa J. Ware

## Abstract

**Background:** The burden of hypertension in South Africa, as well as the successive cardiovascular morbidity and mortality is increasing. Hypertension presents a risk for subsequent cognitive impairment with age. This study sought to determine the association between blood pressure, arterial stiffness, using pulse wave velocity and pulse amplification pressure, and cognitive function in younger and older adults from a 30yr old urban South African birth cohort study.

**Methods:** We conducted a cross-sectional study among n=93 index children (now age 29yr) and their mothers (all women). We collected peripheral and central blood pressure (BP) variables, and conducted a cognitive assessment using the Montreal Cognitive Assessment (MoCA) instrument and analysed the association of BP variables with global cognitive tests and specific domains.

**Results:** Forty percent of the pooled sample had low MoCA total scores, and 32% of the total sample had hypertension. No associations were found in the regression analysis between BP related variables and total MoCA scores. Also, no associations were found between peripheral and central BP variables with individual cognitive domains when stratified by age. A significant relationship was found between mean pressure and low visual perception (i.e. the ability to interpret information that is seen and give it meaning; p=0.02).

**Conclusion:** Central mean pressure is associated with low visual perception domain among black women. These findings add to the growing evidence which suggests that central BP variables are important to explore as exposure proxies for studying the association of BP and cognitive decline especially at mid-life.

## Introduction

Raised blood pressure (BP) and hypertension are well-known modifiable risk factors for the global burden of disease [1], with lowering of BP substantially reducing cardiovascular (CV) morbidity and mortality [2]. Arterial stiffness, the loss of elasticity in large [aorta] and medium [carotid / femoral] arteries, is associated with hypertension (in an indirect feedback loop), and in turn, both hypertension and arterial stiffness are independently associated with cerebrovascular pathogenesis [10]. The burden of hypertension in South Africa (and the successive CV morbidity and mortality) is increasing [3]. The interplay between the mentioned CV and cerebrovascular risk factors also presents a risk for subsequent age-related cognitive impairment [4]–[6] through factors such as wall thickening, luminal narrowing and inflammatory response resulting in atherosclerosis and/or arteriosclerosis [7]–[9]. A recent systematic review by Forte *et al*. (2020) conducted across 50 studies and including data from 107,405 participants (aged 20-93 years) showed that, in individuals without dementia or stroke, higher BP was associated with a higher risk of cognitive decline [10]. Another recent systematic review by Ou *et al*. (2020) reported that this relationship is stronger in mid-life compared to older adults and in those with a systolic BP (SBP) above 130 mmHg [11]. Subgroup analysis however, revealed that the association between hypertension and dementia in older adults was stronger in black adults than in white, Asian or mixed populations. Of note, from the 209 included studies, less than 15 (7%) included Africans and almost all of these were resident in North America or the Caribbean.

Studies in South Africa have shown that early vascular ageing, indicated by increased pulse wave velocity (PWV), can be detected in children from as early as 6 years of age indicating a threat later in life [12]. Furthermore, prevalence of paediatric stage1 BP (≥95^th^ percentile) in Central, Eastern and Southern Africa are approximately twice that of the United States [13], [14]. The higher levels of hypertension observed in black adults, regardless of nationality or country of residence are well known [15]–[17], with the subsequent treatment challenges in low-resource settings being well-described [12], [18].

However, there is less evidence for the prevalence of cognitive impairment and dementia across Africa among young adults [19], potentially due in part to continuing stigmatisation of people living with dementia or poor mental health [20], [21]. Despite calls for more data to raise awareness, promote help-seeking behaviour and engender political commitment to the provision of health services [18], [22] evidence remains scarce. This is especially the case for younger and mid-life adults with elevated BP, where mild cognitive impairment (MCI) may be present [11], [23], and who may benefit from more intensive BP lowering and cognitive interventions [24], [25]. This study therefore sought to determine the association between BP, arterial stiffness and cognitive function in younger and older adults from an urban South African cohort.

## Methodology

### Sampling

In 1990, the longitudinal Birth-to-Twenty Plus (BT20+) study recruited 3273 mother-infant pairs at birth in Soweto, South Africa as previously described [26]. The study was designed to observe young childhood and adolescent growth, development and overall health following the transition to democracy in the Republic of South Africa.

In this study conducted 29 years later, we used cross-sectional data from 156 black female cohort participants (index children and their biological mothers) that attended a CV assessment between August 2019 and March 2020. The assessment was designed to investigate how CV health transmits between generations within the cohort, as such biological mothers and their children were invited to take part. Participants were not invited to participate if: they had apparent physical, mental, or congenital abnormalities. In this analysis participants who reported depression or currently taking anti-depressants were excluded from analysis as depression may cause cognitive deficits [27]. We conducted the study in line with the ethical principles of the Declaration of Helsinki [28] and obtained approval from the Human Research Ethics Committee (Medical) of the University of the Witwatersrand (South Africa) (Ref: M190263). All participants were fully informed about the objectives of the study and written informed consent from each adult participant was obtained.

### Cognitive assessment

We used the Montreal Cognitive Assessment basic (MoCA-B) method, which is designed for participants who may have low levels of education or are illiterate [29]. The test has six cognitive domains: executive functioning, visual perception, verbal fluency, attention, orientation and memory [30]. As this was a non-clinical sample and the aim was to assess the associations between hypertension or elevated BP and MoCA score, we did not conduct further diagnostic screening (cognitive function complaints or disorders). For the score on each domain, the MoCA scoring system was used, for visual perception the scores ranged from 3 to 0 points, and for verbal fluency the points ranged between 2 to 0 points [30].

### Vascular and other measures

Peripheral BP measures were performed using the SphygmoCor XCEL device (AtCor Medical Pty. Ltd., Naperville, USA). Blood pressure readings were recorded on the right upper arm with appropriately sized cuffs. Systolic and diastolic BP (SBP, DBP) were captured from each measurement. Mean arterial pressure (MAP) was subsequently calculated using the formula [SBP+2(DBP)]/3 and pulse pressure was calculated using the formula (SBP–DBP). Augmentation index and central BP readings were then derived by using in-built generalised transfer functions. Additionally, pulse pressure amplification [(brachial pulse pressure / central pulse pressure) *100] was calculated. To obtain the pulse wave velocity (PWV) we used applanation tonometry (high-fidelity micromanometer; Millar Instruments) in conjunction with a partially inflated BP cuff over the right upper thigh to obtain the carotid and femoral pulses simultaneously. The direct distance method was used where 80% of the distance between the carotid and femoral artery was calculated after which the carotid-femoral PWV was estimated along the descending thoracic abdominal aorta [31]. Two measures were taken with a 2-minute resting interval. In the event that the first and second measure differed by more than 0.5 m/s, a third measure was recorded and checked to be within a 0.5 m/s limit with the first or second measure. The average of two measures with ≤ 0.5 m/s variation was used for analysis. For quality control, the PWV measures were repeated if they were not recorded as sufficient quality based on an automated operator index [32].

All anthropometric procedures were performed in triplicate according to specific guidelines and standard protocols [33]. Height was measured with a wall-mounted stadiometer to the nearest 0.01 cm (Holtain, United Kingdom) and weight to the nearest 0.01 kg (Omron calibrated digital scale). Body mass index (BMI) [weight (kg) / square height (m^2^)] of each participant was subsequently calculated. Sociodemographics (age, household information, socio-economic status, education [year of schooling completed]) and medical history (hypertension, history of stroke, diabetes, and depression) were collected through questionnaires. In addition, health behaviour data related to vascular health was collected using the WHO-AUDIT questionnaire (alcohol use), and the Global Adult Tobacco Survey (tobacco use and exposure) [34].

### Data analysis

Only participants with complete cardiovascular and cognitive assessment data were included in the statistical analyses. A score less than 26 was considered a low score as this has previously been classified as mild cognitive impairment (MCI) [29]. Hypertension was classified as a BP ≥140/90 mmHg or being on blood pressure lowering medication. Normality checks were conducted for all data used in this study and there were no departures from normality. For comparisons between the groups (normal or low domain score) we used chi-square (for categorical data) and t-tests (for continuous data) to assess the difference between means. We used ANCOVA to control for MAP in comparing the means of PWV for the two cognitive domains. Additionally, logistic regression models were constructed to determine the association between BP indices and the scores of the MoCA sub-tests/domains. For all statistical analyses, STATA version 15 (Stata Corp, 2017) was used [35].

## Results

Of 156 participants attending the assessment, *n*= 93 participants comprising *n*=46 of the mothers, and *n*=47 of the index children (now age 29 years) had complete data for both the cognitive and cardiovascular assessments. The major reasons for incomplete data were inability to attend all data collection visits due to national lockdowns initiated as a result of the COVID-19 pandemic (*n*=43) or a mid-arm circumference greater than the available blood pressure cuff sizes (*n*=20). In this study, we present pooled (index children and their mothers) analysis results. The characteristics of the women stratified by normal or low MoCA score are presented in **Table 1**. In terms of the whole sample enrolled in the study the mean age (±SD) of the participants in the total sample was 41.8±13 years. Fifty-one of the participants had a National Senior Certificate (NSC) or Matric, and the average BMI was 33.1 ± 8.7 kg/m^2^. Regarding medical history, 32% had ever been diagnosed with hypertension, and 4% had ever been diagnosed with type 2 diabetes (T2DM).

**Table 1.** Socio-demographic, medical history self-reports and behavioural characteristics of the sample in athe study population stratified by normal and low MoCA scores.

|  | Normal MoCA score | Low MoCA score |
| --- | --- | --- |

the sample in athe study population stratified by normal and low MoCA scores.
| **Sample characteristics** | **Mean (SD) /n (%)**<br>**(n=56)** | **Mean (SD) / n(%)**<br>**(n=37)** | **p-value** |
| --- | --- | --- | --- |
| **Age** | 37.7 (12.3) | 48.6 (13.1) | <0.01‡ |
| **Year of schooling completed** |  |  | 0.20 |
| Primary school | 2 (3.5) | 3 (8.1) |  |
| Completed high school (<12 yrs) | 19 (34) | 18(48.7) |  |
| Matric | 35 (62.5) | 30(43.2) |  |
| **Body mass index (kg/m<sup>2</sup>)** |  |  | 0.70 |
| Normal | 10 (17.8) | 6 (16.2) |  |
| Overweight | 1 (1.8) | 0 (0) |  |
| Obese | 45 (80.4) | 31 (83.8) |  |
| **Hypertension, n (%)** |  |  | <0.05† |
| Yes | 13 (23.2) | 17 (46.0) |  |
| No | 43 (76.8) | 20 (54.0) |  |
| **History of stroke** |  |  | 0.15 |
| Yes | 1 (1.78) | 3 (8.1) |  |
| No | 54 (96.4) | 34 (91.1) |  |
| **Diabetes, n (%)** |  |  | 0.54 |
| Yes | 3 (5.0) | 1 (2.7) |  |
| No | 53 (95.0) | 36 (97.3) |  |
| **Currently on depression medication n (%)** |  |  | 0.40 |
| Yes | 1 (1.78) | 0 (0) |  |
| No | 55 (98.2) | 37 (100) |  |
| **Current/past smoker n (%)** |  |  | 0.45 |
| Daily | 3 (5.35) | 3 (8.1) |  |
| Less than daily | 2 (3.6) | 0 (0) |  |
| Not at all | 51 (91.9) | 34 (91.9) |  |
| **Alcohol use, n (%)** |  |  |  |
| Never | 19 (33.9) | 22 (59.4) | 0.02† |
| Monthly or less | 27 (48.2) | 10 (27.0) |  |
| 2 to 4 times a month | 10 (17.8) | 3 (8.1) |  |
| 2 to 3 times a week | 0 (0) | 2 (5.4) |  |
| **MoCA Total Score** | 28.0 (1.02) | 24.0 (1.97) |  |

Note: n – number of participants.

Forty percent of the pooled sample was classified as having a low MoCA score (i.e., score < 26) (i.e., score < 26). The prevalence of low MoCA scores, when stratified by age in the two age groups, was 29% in older adults (age 48 to 68 years) and 10.7% in younger adults (age 29 years). The mean BP levels between normal and low visual perception, and between the normal and low language domain are presented in **Table 2**. The mean differences of central pulse pressure, central augmentation pressure, pulse pressure amplification (PPA) between participants with normal and low visual perception scores were significant (*p*< 0.05). The mean differences of PWV between participants with normal and low verbal fluency and visual perception were significant: (*p*< 0.01) and (*p*< 0.01) respectively, when we controlled for mean arterial pressure.

**Table 2.** Mean blood pressure levels classified by normal and low visuospatial perception and verbal fluency (*n*=93).

| Index | Visual perception |  | Verbal fluency |  |
| --- | --- | --- | --- | --- |
| | Normal<br>(Mean ± SD) | Low<br>(Mean ± SD) | Normal<br>(Mean ± SD) | Low<br>(Mean ± SD) |
| Average brachial SBP (mmHg) | 124 ± 17.0 | 129 ± 20.0 | 125 ± 17.11 | 129 ± 20.4 |
| Average brachial DBP (mmHg) | 77 ± 11.0 | 77 ± 8.0 | 75 ± 11.0 | 78 ± 8.5 |
| Average brachial PP (mmHg) | 38 ± 14.0 | 38 ± 10.0 | 38 ± 14.0 | 38 ± 10.0 |
| Average central SBP (mmHg) | 115 ± 16.1 | 120 ± 20.0 | 114 ± 16.1 | 120 ± 20.0 |
| Average central DBP (mmHg) | 77 ± 11.0 | 78 ± 9.0 | 77 ± 11.0 | 79 ± 9.0 |
| Central PP (mmHg) | 37 ± 9.0† | 42 ± 13.0† | 37 ± 10.0 | 42 ± 13.0 |
| Central Mean pressure (mmHg) | 93 ± 12.0 | 93 ± 12.0 | 93 ± 13.0 | 94 ± 12 |
| Central AP | 12 ± 6.5† | 16 ± 9.2† | 12 ± 6.5 | 16 ± 9.2 |
| Central AI | 31 ± 12.0 | 35 ± 13.3 | 31 ± 12.0 | 35 ± 13.4 |
| PPA | 103 ± 32.4 | 94 ± 26.0 | 103 ± 32.4 | 93 ± 26.0 |
| PWV (m/s) | 23.4 ± 11.1† | 27.0 ± 13.0† | 19.1 ± 9.0‡ | 27.0 ± 12.0‡ |

**Note:** We conducted independent t-tests on the central and peripheral blood pressure measures for the normal vs. low visual perception and verbal fluency. We conducted ANCOVA for pulse wave velocity for both test domains to control for mean arterial pressure. Values are arithmetic mean ± standard deviation.
Abbreviations: *SD* – standard deviation, *SBP* – systolic blood pressure, *DBP* – diastolic blood pressure; *PP* – pulse pressure; *AP* – augmentation pressure; *AI* – augmentation index; *PPA* – pulse pressure amplification; *PWV* – pulse wave velocity.
Significant results are noted below.
† $p < 0.05$ , ‡ $p < 0.01$ .

When individual cognitive domains were analysed using logistic regression (adjusted for age, household assets, BMI and MAP), only central mean pressure (OR: 1.47; 95% CI: 1.06-2.02, *p* = 0.02) was significantly associated with low visual perception as presented in **Figure 1**. No associations were found between other BP indices and low verbal fluency. We additionally assessed if associations presented between all BP indices and the total MoCA score (normal/low), however, no significant associations were found.

**Figure 1:**
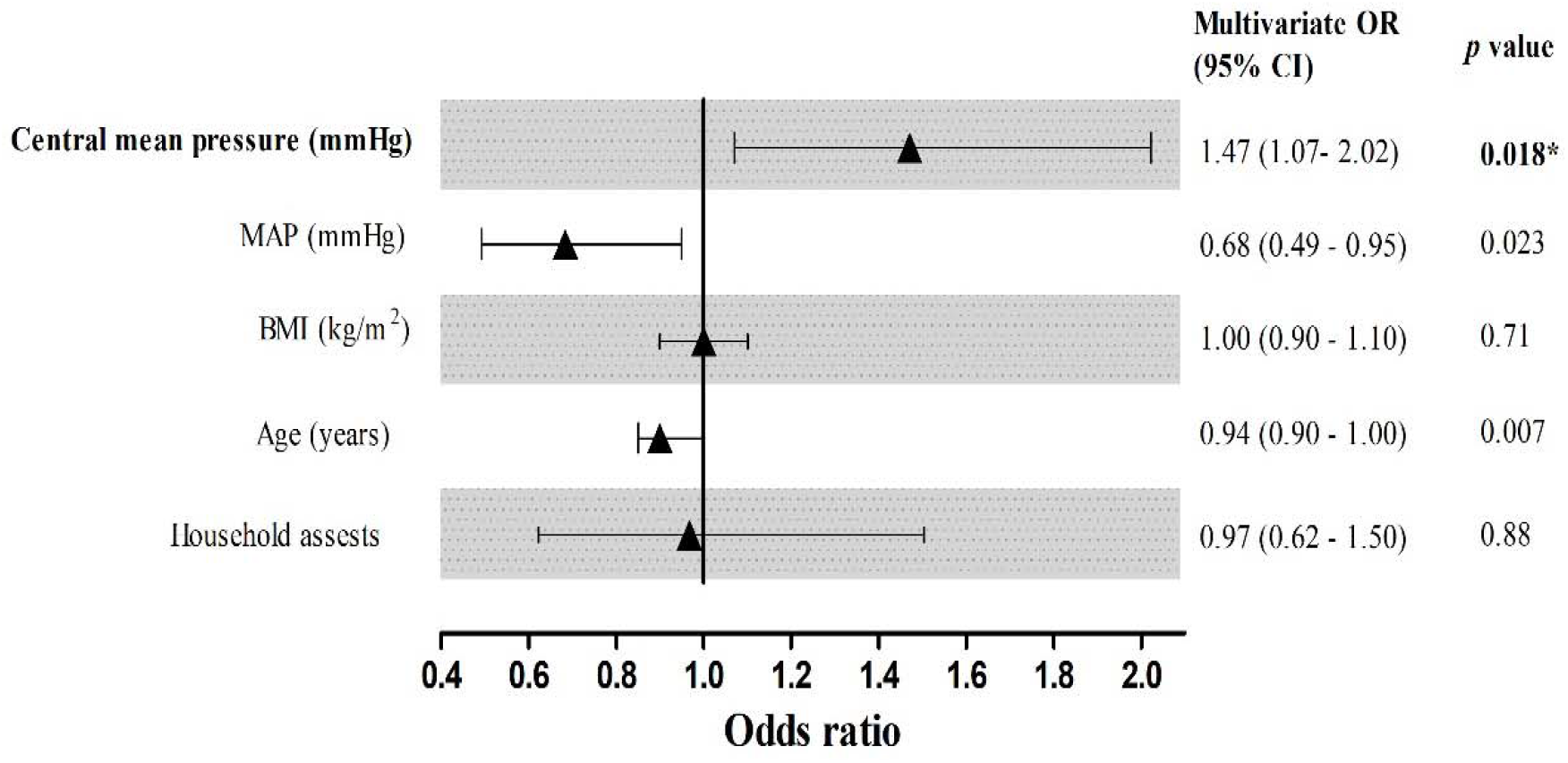
Logistic regressions showing the association between central mean pressure and low visual perception score domains (adjusted for mean arterial pressure (MAP), body mass index (BMI), Age and household assets) (*n*=93).

## Discussion

In this present exploratory study, we found that the lower MoCA total scores in older than in younger adults. In the pooled sample, women who scored low in the visual perception domain had significantly higher central pulse pressure, central augmentation index, and pulse wave velocity. Women with low verbal fluency scores also had a significantly higher pulse wave velocity. When we adjusted for age, BMI and socioeconomic status, only low scores in the visual perception domain were associated with central mean pressure across the sample.

A previous study found an association between central pulse pressure and mild cognitive impairment but that study was conducted in a sample of participants who were older than 60 years of age [36]. A recent study conducted by Suleman *et al*. (2017) that included adults who were 50 years of age did not find an association between pulse pressure and mild cognitive impairment but an association with other central variables (i.e. central augmentation index and pressure pulse amplification). This growing evidence for an association between central blood pressure variables and mild cognitive impairment or lowered cognitive scores highlights the importance of studying these blood pressure variables in non-clinical populations as exposure proxies to domain-specific cognitive decline (i.e. executive function, verbal fluency, attention, visual perception, memory and global cognition). It is possible that we did not see a significant association in our study for other central blood pressure variables (aside from central mean pressure) or pulse wave velocity and lowered cognitive domain scores after adjustment for confounders, because of the size of our sample, and the fact that we included participants who were significantly younger, i.e. who were not quite at midlife yet (<35 years old).

The fact that participants with low cognitive domain scores also had higher means for central pulse pressure, central augmentation index and PWV is noteworthy as literature shows that these variables are indicators of CVD risk or subclinical issues [6], [37], [38]. Also, the fact that there was a significant number of participants with hypertension is concerning as research suggests that BP is a modifiable risk factor that can cause neuropathological changes before the onset of vascular dementia (via atherosclerosis, arteriolosclerosis and cardiac events) [7]. There is also growing evidence that mid-life hypertension is linked to executive function and global cognition decline, suggesting that our younger sample with hypertension are at high risk [11], [12], [16], [17]. The evidence on BP-related factors and their association with cognitive decline or early onset thereof is important for the public health system of South Africa to inform screening and health promotion so that modifiable risk factors such as hypertension can be addressed to prevent adverse health outcomes later in life. Prevention is particularly important because the healthcare system of South Africa is already burdened and does not yet have a comprehensive plan to treat dementia.

We must interpret this study within the context of its strengths and limitations. Similar to a previously reported study, this work contributes to the limited body of knowledge concerning the use of the MoCA assessment for screening mild cognitive impairment or low cognitive score performance in a comprehensive enough fashion (as a variety of cognitive domains are covered) in population-based studies [6], [11], [37]–[39]. Another strength is that this study adds to the African literature and knowledge among young females as research increasingly shows that this may be an important point in the lifespan where early indicators of cognitive decline can be identified, and where interventions can be affected to prevent adverse cognitive outcomes later in life. This study is not without limitations, we conducted our investigations on a small cross-sectional sample thus the findings of which may have led to sample power restrictions. The study being cross-sectional means that we can only infer associations between BP-related factors and low MoCA score and not causality. We also, did not include males in this sub-sample such that we do not know what the association between blood pressure, and arterial stiffness markers looks like in young South African males.

This study was conducted due to increasing evidence highlighting the association of CVD risk factors and cognitive impairment in early life rather than later. It has also been proposed that cardiovascular risk factors (i.e. BP or arterial stiffness) gradually accumulate sub-clinically, thus, the identification of these risk factors in middle-life may further initiate the development of specific guidelines and intervention strategies in preventing and reversing modifiable risk factors that may lead to cognitive impairment in later life.

## Data Availability

All data will be available upon request 3 months after formal publication of this work.

## Appendix 1

**Table 3.** Logistic regression showing the association between blood pressure indices and MoCA cognitive function domains (*n*=93)^a^

| Index | Association with low visual perception scores |  | Association with low verbal fluency scores |  |
| --- | --- | --- | --- | --- |
|  | OR (95% CI) | *p-value* | OR (95% CI) | *p-value* |
| Average brachial SBP (mmHg) | 0.98 (0.89, 1.07) | 0.70 | 1.06 (0.97, 1.16) | 0.19 |
| Average brachial DBP (mmHg) | 1.03(0.85, 1.23) | 0.76 | 0.89(0.74, 1.07) | 0.21 |
| Average brachial PP (mmHg) | 1.03 (1.00, 1.08) | 0.18 | 0.98(0.94, 1.04) | 0.68 |
| Average central SBP (mmHg) | 0.95 (0.86, 1.05) | 0.38 | 1.07(0.97, 1.19) | 0.17 |
| Average central DBP (mmHg) | 1.04 (0.87, 1.25) | 0.64 | 0.88(0.74, 1.05) | 0.16 |
| Average central PP (mmHg) | 0.97(0.90, 1.04) | 0.45 | 1.05 (0.98, 1.12) | 0.15 |
| Central mean pressure(mmHg) | 1.47 (1.06, 2.02) | 0.02* | 0.93 (0.73, 1.18) | 0.57 |
| Central AP | 0.97 (0.88, 1.06) | 0.53 | 1.05 (0.96, 1.15) | 0.25 |
| Central AI | 1.01 (0.95, 1.06) | 0.72 | 1.03 (0.98, 1.08) | 0.18 |
| PPA | 1.03 (0.96,1.11) | 0.30 | 0.96(0.91,1.01) | 0.16 |
| PWV (m/s) | 1.00(0.95, 1.05) | 0.89 | 0.95(0.90,1.00) | 0.07 |

**Note:** Covariates used in the model include age, BMI and household assets.
Abbreviations: *n* – number of participants, OR – odds ratio; CI – confidence interval; *SBP* – systolic blood pressure, *DBP* – diastolic blood pressure; *PP* – pulse pressure; *AP* – augmentation pressure; *AI* – augmentation index; *PPA* – pulse pressure amplification; *PWV* – pulse wave velocity.
Significant results are noted below.
\* $p < 0.05$ .
<sup>a</sup>We also conducted analysis where we stratified by age categories (29y) and (43 to 68y), no significant associations were shown between BP indices and specific cognitive function domains. Also, no significant associations were found between the analysis of BP indices MoCA global score (yes/no). It is important to highlight that for the PWV measure the sample with which we conducted logistic regression was $n= 83$ .

